# Retrospective safety testing of the CT Clock method for identifying treatment-eligible patients with ischaemic stroke of unknown onset time: Study Protocol

**DOI:** 10.64898/2026.05.01.26352219

**Authors:** Grant Mair, Francesca Chappell

## Abstract

More than 100,000 people in the UK have an ischaemic stroke every year. This means a blood clot blocks an artery supplying blood to their brain. Patients with ischaemic stroke often have sudden weakness affecting half of their body or face, and problems with speech, although other stroke symptoms are possible.

Treatment with a clot-busting drug (thrombolysis) can reduce the amount of disability and death caused by ischaemic stroke by restoring the blood supply to the brain. Thrombolysis usually needs to be given within four and a half hours of stroke occurring. Unfortunately, nearly three quarters of patients with ischaemic stroke arrive in hospital later than this, or it is not clear when their stroke started. For these patients, it may still be possible to treat them with thrombolysis if the hospital can provide an additional advanced type of brain scan. However, many hospitals in the UK and worldwide cannot offer this advanced scan to patients with stroke, particularly hospitals that are not in major cities or developed nations. Patients arriving at these hospitals therefore do not currently have the same access to effective treatment for stroke.

We have developed a simple method for identifying which patients can be given thrombolysis even if they arrive at hospital later than four and a half hours, or where there is uncertainty about when their stroke started. Our method does not require any additional or advanced imaging, only the standard computed tomography (CT or CAT) scan that all patients with stroke get when they arrive at hospital. Our method is called the CT Clock. We ask doctors to look for stroke changes indicating ischaemic stroke in the brain on CT. If they find these stroke changes, they measure them compared to normal brain. If the stroke changes on CT are minor (less than 20% darker than normal brain), or the scan appears normal despite quite severe stroke symptoms, we would consider these patients suitable for treatment with thrombolysis.

This study proposes to test the safety of our CT Clock method in an analysis of existing NHS data from patients who have previously been treated with thrombolysis. We aim to show that when doctors who provide stroke care use our CT Clock method to identify suitable patients for treatment with thrombolysis, it is safe for these patients.

If successful, this study will allow us to plan for and deliver future clinical testing where we would use the CT Clock to identify patients for treatment with thrombolysis in hospitals where advanced imaging for stroke is unavailable. Successful clinical testing is needed before our method can be used routinely in hospitals around the world.

**Data Study Protocol:** 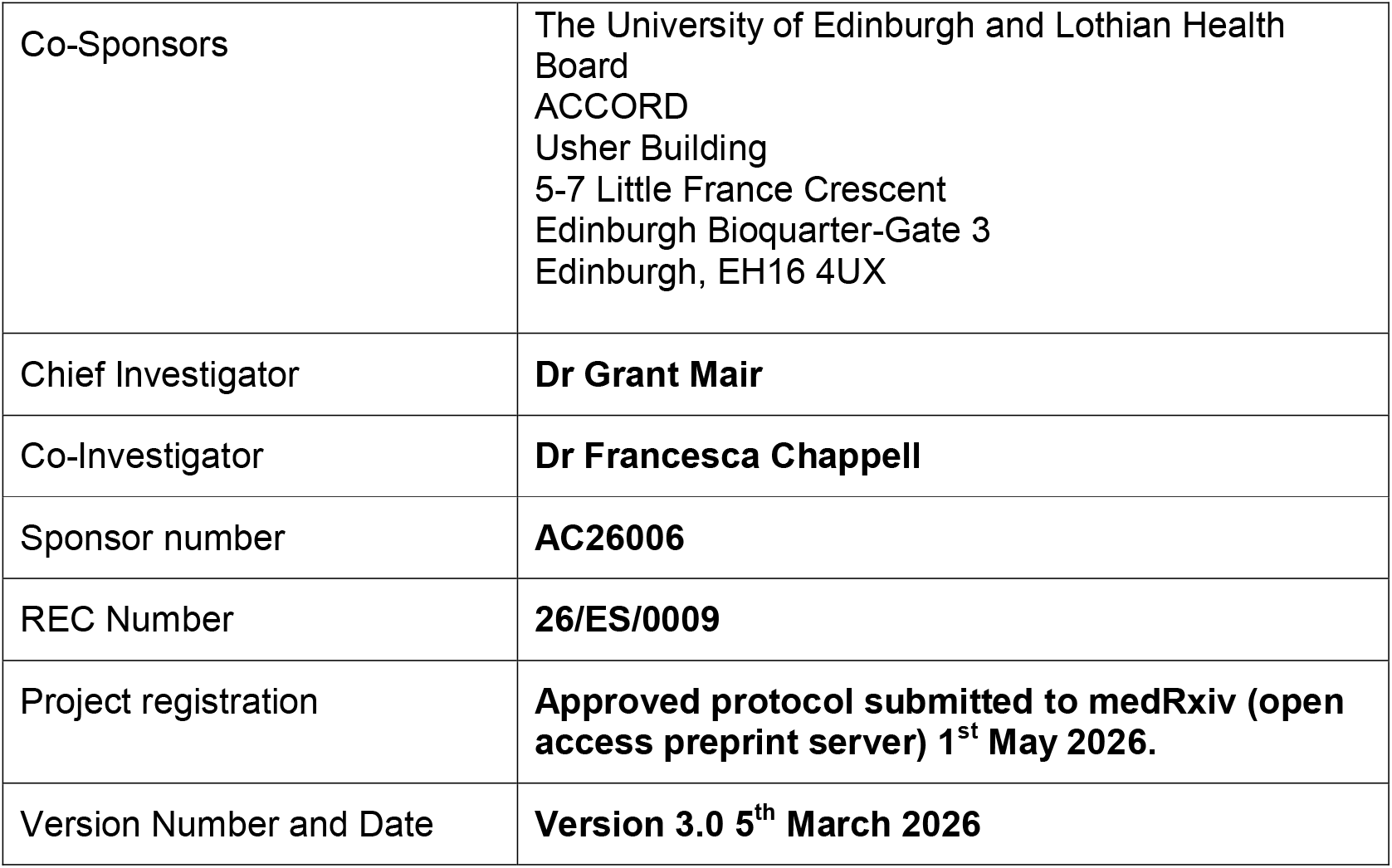

## 1 INTRODUCTION

### 1.1 BACKGROUND

Stroke occurs following disruption of blood supply to the brain causing loss of neurological function. Death rate is high, while survivors are often left disabled. In 2024, there were over 100,000 acute stroke presentations to hospitals in the UK.^1,2^ Most are ischaemic (87%) when an artery supplying the brain is blocked by blood clot, while haemorrhagic stroke (12%) refers to bleeding into the brain. Brain imaging is needed to differentiate ischaemic from haemorrhagic stroke. In the UK, standard imaging is non-enhanced CT, while MRI and contrast-enhanced CT are collectively termed advanced imaging.

Two treatments can restore arterial blood supply in ischaemic stroke. Thrombolysis is the intravenous injection of an agent to break down the occluding blood clot. Thrombectomy involves manual removal of the occluding blood clot via a catheter. Both treatments are most effective when deployed rapidly. European licensing for thrombolysis requires treatment within 4.5 hours of symptom onset, while thrombectomy is routinely offered within 6 hours.^3,4^

Unfortunately, most patients with ischaemic stroke present to hospital later than 4.5 hours (44%) or have an unknown time of symptom onset (30%); this accounts for ∼74,000 patients annually in the UK.^1,2^ For these patients, clinical trials have shown that advanced imaging can differentiate reversible brain ischaemia from irreversible infarct, allowing some patients to be safely and effectively treated with thrombolysis up to 9 hours after symptom onset, or when onset time is unknown.^5,6^ However, around 50% of UK centres have no or limited access to advanced imaging for acute stroke.^1^

We have developed the CT Clock to help bridge this gap.

#### CT Clock method

As ischaemic stroke progresses, injured brain becomes more pronounced with greater oedema. Water appears black on CT, so this appears as tissue darkening, indicating a decrease in CT attenuation, Figure 1.

**Figure 1.**
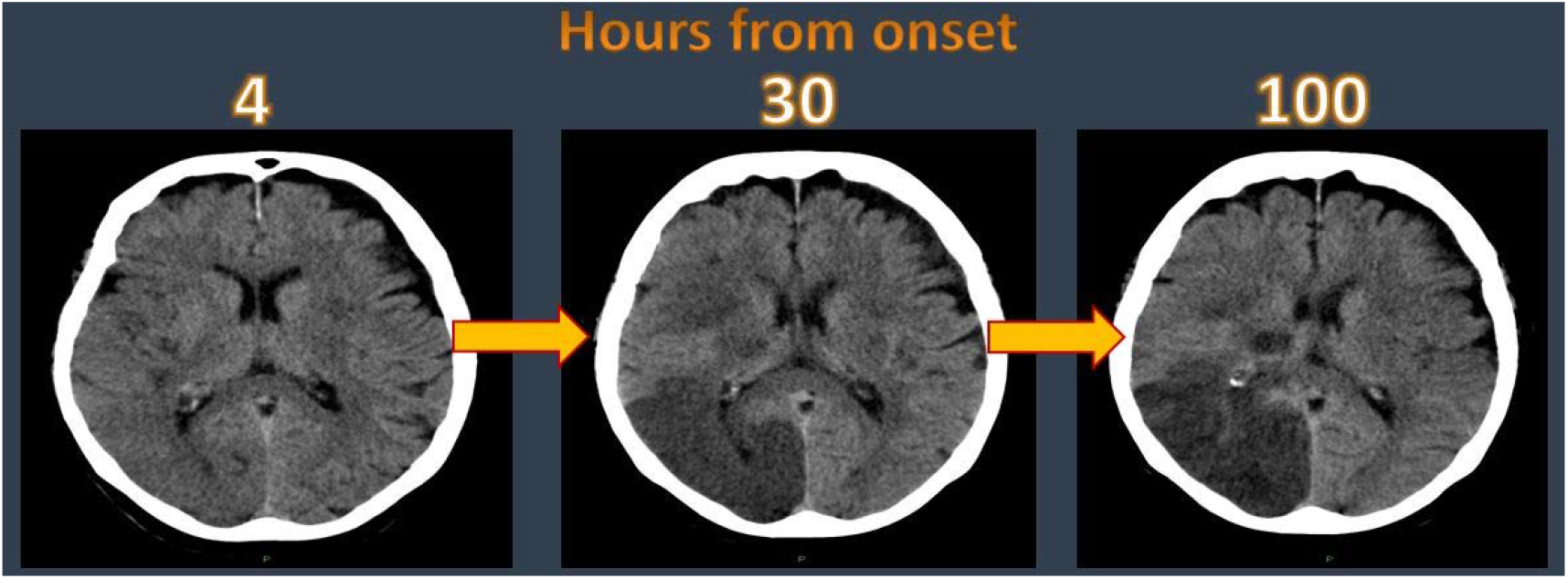
CT attenuation of ischaemic brain with time.

Having captured the rate of CT attenuation change with time in a large cohort,^7^ we can now use CT attenuation to estimate the time of ischaemic stroke onset, and from comparing CT attenuation with concurrently acquired CT perfusion,^8^ we can estimate whether injured brain tissue remains viable. These relationships underpin the CT Clock method.

The CT Clock is designed for wide applicability. It requires only non-enhanced CT and existing image viewing software. Users identify the ischaemic brain lesion and manually measure CT attenuation in injured and contralateral normal brain to derive an attenuation ratio, Figure 2. We use attenuation ratio thresholds to predict when stroke onset time is under 4.5 hours and when brain tissue is viable.

**Figure 2.**
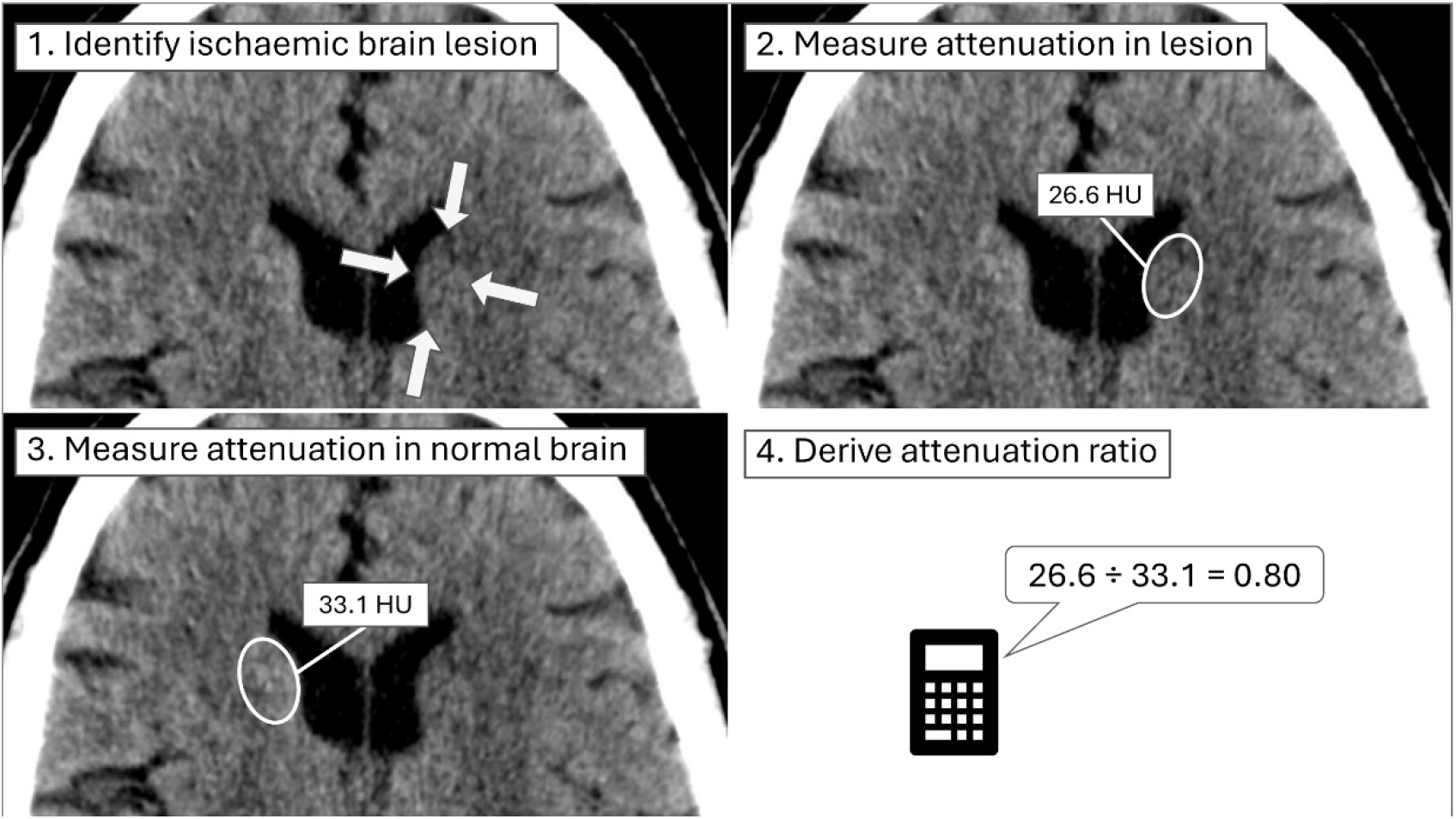
CT Clock method.

### CT Clock development to date

#### CT Clock for estimating the time from stroke onset

We plotted CT attenuation ratio against time for 342 CT scans from a cohort of 200 representative patients from a large clinical trial (The Third International Stroke Trial, IST-3). We used the association between these two variables to determine that an attenuation ratio greater than 0.82 is highly predictive of a stroke onset to CT time less than 4.5 hours (i.e., within the time limit for thrombolysis with alteplase). At this threshold, the CT Clock is 97% sensitive and 83% specific for correctly classifying patients who are within 4.5 hours of stroke symptom onset.^7^

#### CT Clock for estimating viability of ischaemic brain tissue

We have also tested associations between the appearances of non-enhanced CT and CT perfusion (CTP) when both are acquired at the same time. In a single centre analysis of 206 ischaemic lesions from 142 consecutive patients with stroke, we compared the CT attenuation ratio with CTP appearances that indicate affected brain tissue is likely to remain viable. In other words, and in the absence of CTP, a CT attenuation ratio greater than 0.87 is 86% sensitive and 91% specific for identifying brain tissue that remains viable, and is therefore most likely to benefit from treatment.^8^

#### CT Clock when no ischaemic lesion is visible

For patients who present to hospital very early after stroke symptom onset, e.g. <1-hour, ischaemic lesions may not yet be visible on CT (i.e., the attenuation ratio is near normal, very close to 1.0). We have therefore developed a method of identifying patients who are likely to have large non-visible ischaemic lesions (as opposed to no ischaemic lesion in a patient with a stroke mimic or a tiny ischaemic lesion that is overlooked). In an analysis of 2,961 participants from IST-3; we identified a clinical-imaging mismatch (National Institutes of Health Stroke Scale, NIHSS >11 but a normal CT) that predicts non-visible ischaemia.^9^

#### CT Clock clinical feasibility study

In a prospective single centre feasibility study, we asked front-line clinicians (stroke clinicians and radiologists) to apply the method during acute stroke care. Ninety-five consultant and trainee clinicians provided 130 CT Clock assessments for 35 patients with ischaemic stroke (median 3 per patient, range 1-7). All attempts to use the CT Clock were successful. Front-line clinicians found it quick and easy to use the method during their normal practice. Most assessments (120/130, 92%) took <5 minutes. On a scale of 1 Easy to 5 Hard, clinicians found it easy to find (median 2) and measure (median 1) ischaemic lesions (n=72). Estimates for stroke onset time and brain tissue viability derived from clinician measurements had acceptable levels of precision (median absolute error 104 minutes), diagnostic accuracy (82% sensitive, 57% specific for predicting stroke onset time <4.5 hours, 63% of assessments correctly predicted the presence of viable brain tissue) and inter-rater agreement (almost perfect for estimating stroke onset time to the nearest minute). Mair et al. *European Society of Neuroradiology Conference* 2024, full paper under review.

### Classifying CT Clock results

In patients with symptoms of ischaemic stroke, we define a favourable CT Clock result as either:

- A visible ischaemic lesion on baseline non-enhanced CT with attenuation ratio >0.82, or
- No visible ischaemic lesion (clinical-imaging mismatch).

We define a non-favourable CT Clock result as a visible ischaemic lesion on baseline non-enhanced CT with attenuation ratio ≤0.82.

### 1.2 RATIONALE FOR STUDY

Ultimately, we wish to use the CT Clock method to identify patients who can be safely and effectively treated with thrombolysis, when presentation to hospital is delayed or stroke onset time is unknown, and advanced imaging is unavailable.

Before definitive clinical testing, we need robust safety data since thrombolysis is not without harm. Around 15% of patients have worsening symptoms after thrombolysis, due to brain haemorrhage, infarct extension or mass effect.^10^ We need to ensure that the rate of neurological deterioration after thrombolysis is not increased among patients selected using CT Clock methodology.

If we can prove our method is safe, we can proceed to prospective clinical testing. Ultimately, our simple and widely applicable method could substantially increase the number of patients treated with thrombolysis globally; for every ten patients treated with thrombolysis, one additional patient will have an excellent outcome.^11^

## 2 STUDY OBJECTIVES

### 2.1 OBJECTIVES

#### 2.1.1 Primary Objective

Assess safety of intravenous thrombolysis given to patients with a favourable CT Clock result.

#### 2.1.2 Secondary Objectives

1. Refine the definition for a favourable CT Clock result.
2. Assess quality of CT Clock results provided by front-line clinicians.

### 2.2 ENDPOINTS

#### 2.2.1 Primary Endpoint

Proportion of patients with early neurological deterioration (END) after thrombolysis in those with favourable and non-favourable CT Clock results.

#### 2.2.2 Secondary Endpoints

1. Proportion of patients with brain haemorrhage or death after thrombolysis in those with favourable and non-favourable CT Clock results.
2. Definition for CT Clock favourable result that is safe and includes the greatest range of patients.
3. Precision and inter-agreement of CT Clock results provided by front-line clinicians.

## 3 STUDY DESIGN

### Overview

Single-centre retrospective analysis of routinely collected non-consented NHS Lothian data from consecutive patients with ischaemic stroke who were treated with thrombolysis. We will classify patients at baseline (before thrombolysis) into those with favourable and non-favourable CT Clock results, then compare short-term post-thrombolysis outcomes between groups.

### Site

Royal Infirmary of Edinburgh (RIE), a comprehensive stroke centre with ∼1500 stroke presentations annually, offering thrombolysis, thrombectomy and high-dependency stroke care.

### Target population

Adult patients treated with thrombolysis for ischaemic stroke over 3+ years (duration will depend on rate of END in the target population).

### 3.1 DATASET

For each patient, we will review existing imaging and non-imaging data relating to the stroke presentation and subsequent in-hospital outcomes

- Imaging at baseline (i.e. acquired soon after presentation to hospital with stroke)
  ∘ Non-enhanced brain CT.
  ∘ CT angiography and CT perfusion*
  ∘ Associated radiology reports including referral history.
- Clinical data at baseline
  ∘ Demographics (age, sex)
  ∘ Stroke severity - NIHSS (National Institutes of Health Stroke Scale)*
  ∘ Stroke subtype - OCSP (Oxford Community Stroke Program Classification)*
  ∘ Elapsed time between stroke onset and baseline CT (in minutes)*
  ∘ Thrombolytic agent (alteplase or tenecteplase)
  ∘ Thrombectomy (successful, yes or no)
- Follow-up imaging relating to same stroke episode (within 7 days)
  ∘ Any further CT or MRI and associated radiology reports*
- Follow-up clinical data (within 7 days)
  ∘ Any neurological deterioration or death.

* May not be available for all patients.

All data will be electronic

- Imaging data will remain within the NHS PACS (Picture Archiving and Communication System) server at all times, viewed as DICOM (Digital Imaging and Communications in Medicine).
- Clinical data will be transcribed from the NHS Lothian electronic health records into the study electronic Case Report Form (eCRF) for storage in secure University of Edinburgh servers via our secure study webpage. Data transfer to the study webpage will be encrypted using https. Transferred clinical data will be stored using a pseudonymisation ID. The study Chief Investigator (CI) and approved co-investigators will transcribe clinical data. As a retrospective analysis, baseline and follow-up clinical data for a single patient can be collected in one sitting.

Transfer of pseudonymised clinical data from the NHS to the University of Edinburgh

- Our study webpage is hosted within secure University of Edinburgh servers. The study webpage provides a portal for data submission to our eCRF which is hosted on the same University of Edinburgh servers.

Imaging analysis within NHS Lothian

- Imaging data will be assessed by the CI and a range of front-line clinicians who routinely review acute stroke imaging. These assessments will only occur within NHS Lothian premises using the existing PACS system.
- Clinical data and results of imaging assessment will be assessed by the CI and study statistician within the University of Edinburgh.

See Figure 3 for data flow through the study.

**Figure 3.**
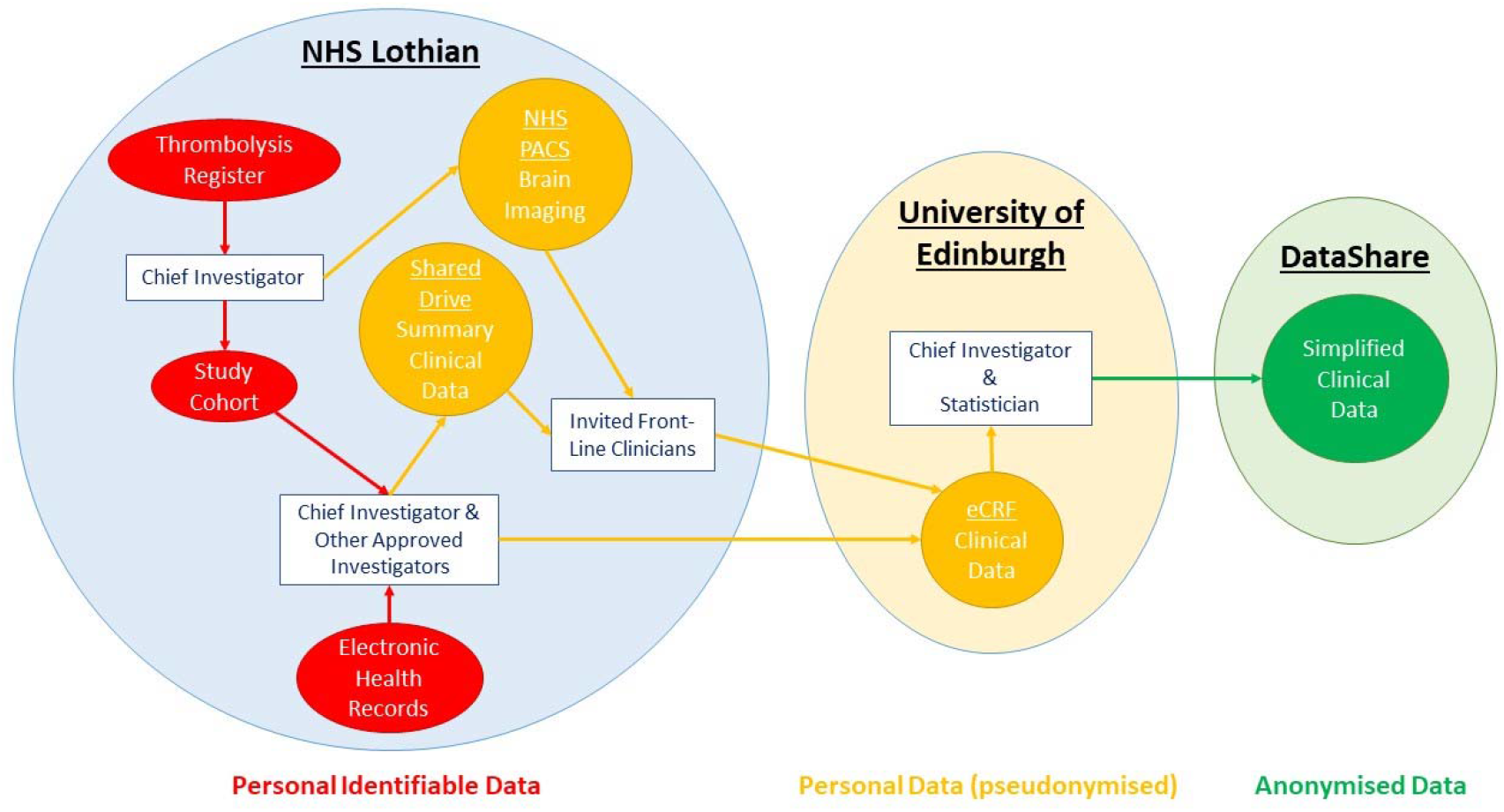
Data flow chart.

### 3.2 SOURCE

All data will be routine clinical data sourced from NHS Lothian.

### Cohort identification within NHS Lothian network

The CI will use an excerpt of the RIE thrombolysis register which includes identifiable details for all patients treated with thrombolysis at RIE (the excerpt will include CHI and date of thrombolysis only for the most recent 5 years). The CI will first review the most recent 12 months of data to ascertain the rates of END and favourable CT Clock results per annum. The total cohort size will then be calculated based on these rates, see 4.1 SAMPLE SIZE CALCULATION. The CI will produce a final list of patients (the study cohort) identified using CHI (Community Health Index) numbers and including the date of thrombolysis treatment.

### Pseudonymisation and data collection

The CI will create a pseudonymisation key from the study cohort list. This will be a separate spreadsheet containing only three columns, the CHI number, an allocated study ID number, and the date of thrombolysis for each patient in the study cohort. The pseudonymisation key will remain within NHS Lothian at all times, within a shared network drive. Access to the pseudonymisation key will be restricted to the CI, and co-investigators with existing access to NHS Lothian data and systems.

The CI will use the pseudonymisation key and thrombolysis date to identify relevant brain imaging at baseline and within 7 days of thrombolysis for each patient in the study cohort. These scans will be added to the Carestream PACS teaching folder labelled using the study ID only. Note, this does not move or otherwise alter imaging, but ensures that when viewed by this route, the data are pseudonymised (all person-identifiable information is removed). The Carestream teaching folder also removes access to the radiology referral history and report. The CI will therefore copy the referral history and radiology report (manually to allow removal of any unexpected personal identifiable data) to a separate spreadsheet labelled using only the study IDs and imaging dates. This spreadsheet will also remain within the NHS Lothian network.

The CI and approved co-investigators will also use the pseudonymisation key to guide collection of clinical data for each patient based on the date of thrombolysis. Only study IDs (not CHI) will be added to the eCRF.

### Analysis

Where the original radiology report identifies an ischaemic brain lesion, invited clinicians will be asked to provide CT Clock assessments from pseudonymised data only. Each clinician will see only a small batch of baseline non-enhanced CT brain scans from within the Carestream teaching folder (e.g. 20, depending on the number of cases with visible lesions and the number of participating clinicians) and a summary of the associated baseline data (age, sex, stroke severity and type, summary of clinical presentation history, text of the original radiology report) on a spreadsheet accessible from a shared drive on the NHS Lothian network (not the same shared drive holding the pseudonymisation key). Clinicians will be asked to follow the CT Clock method (Figure 2; similar instructions will be provided to clinicians) to obtain CT attenuation measurements within the ischaemic lesion (guided by the CT report) and corresponding normal brain. Clinicians will submit their results to the eCRF, which is hosted within the University of Edinburgh. To assess agreement between measurements provided by different clinicians, 20% of CT Clock assessments will overlap. However, secondary assessments will only be used for testing inter-rater agreement. CT Clock assessments will only occur within NHS Lothian premises for staff who already have access to NHS PACS.

The CI and study statistician will have access to all pseudonymised data for analysis within the University of Edinburgh.

See Figure 3 for data flow through the study.

### 3.3 INCLUSION CRITERIA

We will include adult patients (no upper age limit) with ischaemic stroke treated with thrombolysis where onset time is both known and unknown. We will include patients who did and did not receive thrombectomy.

### 3.4 EXCLUSION CRITERIA

We will exclude patients with stroke symptoms not caused by ischaemia or where treatment with thrombolysis was not completed.

### 3.5 CONFIDENTIALITY & CONSENT

As a retrospective analysis of existing NHS patient data extending back several years, it will not be practical and may not be possible to acquire consent from participants (given the severity of stroke, many will not have survived or have capacity to provide informed consent). As such, use of these data for research will require approval from an independent Research Ethics Committee, and from the local NHS Caldicott Guardian, amongst others (see section 3.7 DATA STATUS - APPROVALS for more details). As the proposed data are from a single Scottish NHS site and where relevant will be pseudonymised before leaving the NHS network, we have been advised by our local research office that PBPP (Public Benefit and Privacy Panel for Health and Social Care) approval will not be required.

As a retrospective analysis, there should be no impact on individuals and there are no procedures for participants. The only potential risk relates to data security. Amelioration of this risk is discussed in detail in our separate Data Management Plan but is also summarised in sections 3.6 to 3.13.

### 3.6 DATA PROTECTION

As a retrospective analysis of existing NHS patient data, we will not be able to inform subjects in advance about how we plan to use their data.

Data protection is paramount, and our use of these data will meet the ‘data protection by design and default’ principles of the UK GDPR legislation at every stage of the Data Information Flow.

Key features of our data handling plan include:

1. Collecting only those data essential for the analysis.
2. Pseudonymisation of all data prior to analysis.
3. Only a few essential individuals will have access to patient identifiable data and only within the NHS network.
4. Access to pseudonymised imaging and clinical data will be limited to invited clinicians and only within the NHS Lothian network.
5. Transfer of pseudonymised patient data (transcribed text only) from the NHS to the University of Edinburgh will be encrypted.
6. At study end, data that can be anonymised (limited to summary text data and data derived from analysis only) will be shared openly, while any data that cannot definitively be anonymised (e.g. dates, copies of referral history and radiology reports) will be destroyed.

The main risk in our study is unauthorised access to electronic pseudonymised data outside of the NHS network. All access via the University network is audited and these networks maintain industry standards for access control. Therefore, these risks are deemed low.

### 3.7 DATA STATUS – APPROVALS

Non-consented pseudonymised data will be transferred between NHS Lothian and the University of Edinburgh.

Use of these data for research will require:

- ACCORD sponsorship
- Approval from an independent Research Ethics Committee
- Approval from the Caldicott Guardian and Research & Development office within NHS Lothian.

### 3.8 DATA STORAGE

All data will be electronic.

Personal identifiable data (including the pseudonymisation key) will not leave the NHS.

Personal data (pseudonymised) will be transferred to servers within the University of Edinburgh for analysis. Specifically, per patient this will include age, sex, stroke severity (NIHSS), stroke type (OCSP), number of minutes between stroke onset and CT, thrombolytic agent used (alteplase or tenecteplase), whether thrombectomy occurred (yes or no) and if this was successful (yes or no), whether haemorrhage was identified on follow-up imaging (yes or no), and if present whether this haemorrhage was likely to be associated with worsening symptoms (yes or no), whether END occurred within 7 days of stroke (yes or no), whether death occurred within 7 days of stroke (yes or no), attenuation measurements acquired from CT by front-line clinicians and CI, front-line clinician confidence in their assessments (1 to 5).

All University of Edinburgh servers are in the UK, and they provide enterprise-class storage with guaranteed backup and resilience. Only the research team will have access to pseudonymised data within the University of Edinburgh.

### 3.9 DATA RETENTION

All data including that stored within the NHS and the University of Edinburgh will be retained for up to 3 years from the end of the project.

Upon publication of the main study paper, the pseudonymisation key will be permanently deleted. At this point therefore, all remaining study data will be anonymised.

After the end of the project, anonymised text data will be simplified and moved to the University of Edinburgh DataShare digital repository for open sharing (no fixed end point). This move will be timed so that corresponding study publication drafts can include relevant data access DOI(s). However, open sharing of the data will be embargoed until the data are anonymised (pseudonymisation key is deleted).

Specifically, for each patient we will openly share the following, where available:

- Demographics (age, sex)
- Stroke severity (NIHSS) and subtype (OCSP)
- Elapsed time between stroke onset and CT (in minutes)
- CT Clock results provided by front-line clinicians (favourable or non-favourable)
- CT Clock result provided by expert (favourable or non-favourable)
- Thrombolytic agent (alteplase or tenecteplase)
- Thrombectomy (successful, yes or no)
- Neurological deterioration (END) within 7 days (yes or no)
- Brain haemorrhage on follow-up imaging (yes or no)

All other study data will then be deleted.

#### 3.10 DISPOSAL OF DATA

Deletion of study data from University of Edinburgh servers will automatically include the backups of these data after 60 days.

Prior to open sharing, text data will be copied to clean spreadsheets without the original study IDs. Spreadsheet rows will be reordered (e.g. by age) and a new ID applied.

### 3.11 EXTERNAL TRANSFER OF DATA

Person identifiable data will not leave the NHS.

Personal (pseudonymised) data will not leave the University of Edinburgh.

Only anonymised (non-imaging) data will be shared openly.

### 3.12 DATA CONTROLLER

The University of Edinburgh and Lothian Health Board are joint data controllers.

### 3.13 DATA BREACHES

Any data breaches will be reported to the University of Edinburgh and NHS Lothian Data Protection Officers who will onward report to the relevant authority according to the appropriate timelines if required.

## 4 STATISTICS AND DATA ANALYSIS

### 4.1 SAMPLE SIZE CALCULATION

To test whether patients with a favourable CT Clock result treated with thrombolysis have similar risk of harm to patients with a non-favourable CT Clock result, we propose non-inferiority testing of post thrombolysis outcomes between these groups.

The most feared complication of thrombolysis is secondary brain haemorrhage causing a worsening of stroke symptoms. So called symptomatic intracranial haemorrhage (SICH) is associated with poorer long-term outcomes after stroke including an increased risk of death. Depending on the definition used, SICH is fortunately very rare (2-6% of cases), including in trials where time of stroke onset is unknown or presentation delayed. ^12-14^ Also, for pragmatic and ethical reasons, SICH is not always confirmed with repeat brain imaging in routine practice. For example, in patients who are palliated. It is therefore difficult to adequately power non-inferiority testing using routine clinical data based on SICH alone. Early neurological deterioration (END) after thrombolysis is a broader term associated with several pathological processes occurring in the days after treatment including SICH, brain swelling, and infarct extension that does not rely on repeat brain imaging for diagnosis. A retrospective study from a large UK stroke centre sought clinical deterioration from a range of electronic health records and found END in 14% of patients, mostly within 7 days of thrombolysis. ^10^ We will use a similar means of identifying END from data at our centre as the primary outcome measure for CT Clock safety testing.

### Sample enrichment

RIE sees ∼1500 patients with stroke per year and 14% of these are thrombolysed (210 per year). If 14% of patients given thrombolysis have END, we can expect 29 cases per year.

To ensure that non-inferiority testing is adequately powered in a manageable sample size, we will increase the rate of END in our sample to 30%. Most of our sample will be patients treated consecutively with thrombolysis (with and without END). We will then enrich this sample with consecutive patients with END (excluding additional thrombolysed patients without END) to increase the END proportion to 30%.

### Power calculation for non-inferiority testing

For CT Clock safety testing, we assume no difference in END rate between groups with favourable and non-favourable CT Clock results. In a cohort with 30% END, if we set a non-inferiority margin at 15%, we estimate (using this app) that 160 patients per arm will have 90% power to assess whether thrombolysis given to patients with a favourable CT Clock result is non-inferior to thrombolysis for patients with a non-favourable CT Clock result.

#### Handling sample uncertainties

There are two uncertainties in our plan:

1. Unknown proportion of patients with favourable and non-favourable CT Clock results, but unlikely to be exactly 50%. Our recent prospective feasibility study of the CT Clock found that 62% (81/130) had a favourable result.
2. Unknown proportion of patients with END.

With a minimum total sample size of 320, we expect our consecutive thrombolysis sample to be at least 12 months in duration (210 patients). From this 12-month sample, we will identify the proportion with a favourable CT Clock result and the proportion with END. These proportions will determine the total sample size and the number of additional cases with END needed to reach 30% enrichment. For example, if 62% have a favourable CT Clock result, we will need a total sample size of 422 to ensure that 160 (38% of 422) have a non-favourable CT Clock result. Meanwhile, if 14% have END, increasing this to 30% of 422 (127 with END) would require approximately 20 months of consecutive thrombolysis cases (343 total, 48 with END) enriched with a further 79 cases with END.

### 4.2 PROPOSED ANALYSES

All cases in our cohort will be assessed using CT Clock methodology (see section 1.1). This will include clinically representative front-line clinician assessment (index testing) and expert assessment (reference testing).

#### Index testing

We will invite a range of front-line clinicians to review subsets of baseline non-enhanced CT (up to 20 cases per individual) and to provide a CT Clock assessment of ischaemic lesions identified in the original radiologist report for the CT.

- Clinicians will be provided with patient demographics (age & sex), excerpts from the radiology referral text (clinical history), stroke severity and subtype information, and the radiologist report for the non-enhanced CT only, masked to follow-up data.
- For CT Clock assessment, clinicians will be asked to find the acute ischaemic brain lesions described in the original radiology report for the same CT, and then to measure the attenuation of this lesion and of contralateral normal brain.
- For inter-rater evaluation, 20% of all clinician-rated cases will overlap with cases rated by other clinicians.
- Where the original radiologist report does not identify an acute ischaemic lesion, CT Clock classification is provided without further review by front-line clinicians (see below).

#### Reference testing

Expert CT Clock assessment of baseline imaging.

- Expert will be provided with the radiology referral text and all baseline imaging (including CT angiography and CT perfusion if available) with the corresponding radiology report, masked to follow-up data.
- Expert will review all CTs regardless of whether the original radiologist report identifies acute ischaemia.

#### We will then classify cases into CT Clock favourable result subgroups

- Favourable – Visible ischaemic lesion with attenuation ratio >0.82 or no visible ischaemic lesion
- Non-favourable – Visible ischaemic lesion with attenuation ratio ≤0.82 Finally, we will compare outcome data by CT Clock subgroups:

We will use non-inferiority statistics to test whether the rate of END after thrombolysis in the favourable and non-favourable CT Clock groups differ. With 30% END in the entire cohort and a non-inferiority margin of 15%, we estimate that 160 patients with favourable and 160 with non-favourable CT Clock results are needed to prove that the rate of END does not differ statistically between groups (see section 4.1 for full power calculation).

#### Primary outcome

- We will compare post-thrombolysis END between groups with favourable and non-favourable CT Clock results (according to front-line clinician assessment) in absolute terms (as a percentage), but also as odds ratios in a logistic regression.

#### Secondary outcomes, we will

- Compare rates of post-thrombolysis brain haemorrhage (any type) and death between groups with favourable and non-favourable CT Clock results (according to front-line clinician assessment).
- Compare rates of END, post-thrombolysis brain haemorrhage (any type) and death between groups with favourable and non-favourable CT Clock results (according to expert assessment).
- Assess inter-rater reliability for CT Clock assessments provided by front-line clinicians.

#### We will conduct sensitivity analyses on our primary outcome for important subgroups

- To ensure that sample enrichment with additional non-consecutively thrombolysed END cases did not skew results, we will conduct a sensitivity analysis on the consecutive cases only.
- There is increasing uptake of tenecteplase as the principal thrombolytic agent used in the NHS (used routinely in RIE since Sep 25). We will perform a sensitivity analysis including only patients treated with tenecteplase.
- Previous testing suggested we may need to exclude patients from CT Clock assessment with non-visible ischaemic lesions and low NIHSS. We will perform sensitivity analyses excluding patients with no visible lesion and low NIHSS defined as ≤4, ≤6 or ≤10.

## 5 OVERSIGHT ARRANGEMENTS

### 5.1 INSPECTION OF RECORDS

Investigators and institutions involved in the study will permit trial related monitoring and audits on behalf of the Sponsor, REC review, and regulatory inspection(s). In the event of audit or monitoring, the Investigator agrees to allow the representatives of the Sponsor direct access to all study records and source documentation. In the event of regulatory inspection, the Investigator agrees to allow inspectors direct access to all study records and source documentation.

### 5.2 GOOD CLINICAL PRACTICE

#### 5.2.1 Ethical Conduct

The study will be conducted in accordance with the principles of the International Conference on Harmonisation Tripartite Guideline for Good Clinical Practice (ICH GCP).

Before the study can commence, all required approvals will be obtained and any conditions of approvals will be met.

### 5.3 INVESTIGATOR RESPONSIBILITIES

The Investigator is responsible for the overall conduct of the study at the site and compliance with the protocol and any protocol amendments. In accordance with the principles of ICH GCP, the following areas listed in this section are also the responsibility of the Investigator. Responsibilities may be delegated to an appropriate member of study site staff. Delegated tasks should be documented on a Delegation Log and signed by all those named on the list prior to undertaking applicable study-related procedures.

#### 5.3.1 Study Site Staff

The Investigator must be familiar with the protocol and the study requirements. It is the Investigator’s responsibility to ensure that all staff assisting with the study are adequately informed about the protocol and their trial related duties.

#### 5.3.2 Data Recording

The Principal Investigator is responsible for the quality of the data recorded at each Investigator Site.

### 5.3.3 Investigator Documentation

The Principal Investigator will ensure that the required documentation is available in local Investigator Site files ISFs.

#### 5.3.4 Training

##### 5.3.4.1 GCP Training

For data studies, all researchers are encouraged to undertake GCP training in order to understand the principles of GCP. However, this is not a mandatory requirement unless deemed so by the Sponsor. GCP training status for all Investigators should be indicated in their respective CVs.

##### 5.3.4.2 Data Protection Training

All University of Edinburgh employed researchers and study staff will complete the Data Protection Training and Data Protection for Research through Learn.

#### 5.3.4.3 Information Security Training

All University of Edinburgh employed researchers, students and study staff will complete the Information Security Essentials modules and will have read the minimum and required reading setting out ground rules to be complied with.

#### 5.3.5 Confidentiality

All evaluation forms, reports, and other records must be identified in a manner designed to maintain participant confidentiality. All records must be kept in a secure storage area with limited access. Clinical information will not be released without the written permission of the participant. The Investigator and study site staff involved with this study may not disclose or use for any purpose other than performance of the study, any data, record, or other unpublished, confidential information disclosed to those individuals for the purpose of the study. Prior written agreement from the Sponsor or its designee must be obtained for the disclosure of any said confidential information to other parties.

#### 5.3.6 Data Protection

All Investigators and study site staff involved with this study must comply with the requirements of the appropriate data protection legislation (including the UK General Data Protection Regulation legislation and Data Protection Act) with regard to the collection, storage, processing and disclosure of personal information.

Access to collated participant data will be restricted to individuals from the research team treating the participants, representatives of the Sponsor and representatives of regulatory authorities.

Computers used to collate the data will have limited access measures via user names and passwords.

Published results will not contain any personal data that could allow identification of individual participants.

Confirm that relevant information security policies and standards will be adhered to and the investigator should consider further protective measures in consultation with IS and with due consideration of IS policies and standards.

## 6 STUDY CONDUCT RESPONSIBILITIES

### 6.1 PROTOCOL AMENDMENTS

Any changes in research activity, except those necessary to remove an apparent, immediate hazard to the participant in the case of an urgent safety measure, must be reviewed and approved by the Chief Investigator.

Proposed amendments will be submitted to a Sponsor representative for review and authorisation before being submitted in writing to the appropriate REC and local R&D for approval prior to participants being enrolled into an amended protocol.

### 6.2 MANAGEMENT OF PROTOCOL NON-COMPLIANCE

Prospective protocol deviations, i.e. protocol waivers, will not be approved by the Sponsor and therefore will not be implemented, except where necessary to eliminate an immediate hazard to study participants. If this necessitates a subsequent protocol amendment, this should be submitted to the REC and local R&D for review and approval if appropriate.

Protocol deviations will be recorded in a protocol deviation log and logs will be submitted to the Sponsor every 3 months. Each protocol violation will be reported to the Sponsor within 3 days of becoming aware of the violation. All protocol deviation logs and violation forms should be emailed to.

Deviations and violations are non-compliance events discovered after the event has occurred. Deviation logs will be maintained for each site in multi-centre studies. An alternative frequency of deviation log submission to the Sponsor may be agreed with the Sponsor in writing.

### 6.3 SERIOUS BREACH REQUIREMENTS

A serious breach is a breach which is likely to affect to a significant degree:

(a) the safety or physical or mental integrity of the participants of the trial; or

(b) the scientific value of the trial.

If a potential serious breach is identified by the Chief investigator, Principal Investigator or delegates, the Sponsor must be notified within 24 hours. It is the responsibility of the Sponsor to assess the impact of the breach on the scientific value of the trial, to determine whether the incident constitutes a serious breach and report to research ethics committees as necessary.

### 6.4 END OF STUDY

The end of study is defined as the last datapoint analysed.

The Investigators or the Sponsor have the right at any time to terminate the study for clinical or administrative reasons.

The end of the study will be reported to the REC, and R&D Office(s) and Sponsor within 90 days, or 15 days if the study is terminated prematurely. End of study notification will be reported to the Sponsor via email to.

A summary report of the study will be provided to the REC and Sponsor within 1 year of the end of the study.

### 6.5 INSURANCE AND INDEMNITY

The Sponsor is responsible for ensuring proper provision has been made for insurance or indemnity to cover their liability and the liability of the Chief Investigator and staff.

The following arrangements are in place to fulfil the Sponsor responsibilities:

The Protocol has been designed by the Chief Investigator and researchers employed by the University and collaborators. The University has insurance in place (which includes no-fault compensation) for negligent harm caused by poor protocol design by the Chief Investigator and researchers employed by the University.

Sites participating in the study will be liable for clinical negligence and other negligent harm to individuals taking part in the study and covered by the duty of care owed to them by the sites concerned. The Sponsor requires individual sites participating in the study to arrange for their own insurance or indemnity in respect of these liabilities.

Sites which are part of the United Kingdom’s National Health Service will have the benefit of NHS Indemnity.

## 7 REPORTING, PUBLICATIONS AND NOTIFICATION OF RESULTS

## Data Availability

After the end of the project, anonymised text data will be moved to the University of Edinburgh DataShare digital repository for open sharing.

## LIST OF ABBREVIATIONS

ACCORD: Academic and Clinical Central Office for Research & Development - Joint office for The University of Edinburgh and Lothian Health Board
CH: Community Health Index
CI: Chief Investigator
CRF: Case Report Form Computed Tomography
DICOM: Digital Imaging and Communications in Medicine Electronic Case Report Form
EMERGE: Emergency Medicine Research Group of Edinburgh
END: Early Neurological Deterioration
GCP: Good Clinical Practice
GDPR: General Data Protection Regulation
IT: Information Technology
MRI: Magnetic Resonance Imaging
NHS: National Health Service
NIHSS: National Institutes of Health Stroke Scale
OCSP: Oxford Community Stroke Program Classification
PACS: Picture Archiving and Communication System
PBPP: Public Benefit and Privacy Panel for Health and Social Care
PI: Principal Investigator
QA: Quality Assurance
REC: Research Ethics Committee
RIE: Royal Infirmary of Edinburgh
SICH: Symptomatic Intracranial Haemorrhage
SMARTIS: Systematic Management, Archiving & Reviewing of Trial Images Service

## 7.1 AUTHORSHIP POLICY

Ownership of the data arising from this study resides with the study team.

